# The Association between Macular Structure and Idiopathic Epiretinal Membrane

**DOI:** 10.1101/2020.12.20.20248565

**Authors:** I-Hsin Ma, Chung-May Yang, Yi-Ting Hsieh

**Author notes:** **Correspondence:** Yi-Ting Hsieh, MD PhD, Department of Ophthalmology, National Taiwan University Hospital, 7 Zhongshan S. Rd., Zhongzheng Dist., Taipei, 10002, Taiwan. **Conflict of Interest:** No conflicting relationship exists for any author.

## Abstract

**Purpose:** This study measured the anatomical characteristics of the macula in fellow eyes of patients with unilateral idiopathic epiretinal membrane (ERM) and compared them with normal control.

**Design:** A retrospective observational study.

**Participants:** A total of of 94 fellow eyes with unilateral idiopathic ERM were gathered as the study group, and their age and gender-matched subjects with no vitreomacular diseases were recruited as the control group.

**Methods:** Macular structure parameters including foveal base width (FBW), central foveolar thickness (CFT), central subfield thickness (CST), area of foveal avascular zone (FAZ), and retinal artery trajectory (RAT) were measured using optical coherence tomography (OCT) and OCT angiography and were compared between two groups.

**Main outcome measures:** Macular structure parameters were compared between the two groups. A wide-based foveal pit was defined as a FBW larger than the mean value plus one standard deviation of the normal population. Regression analysis was performed to evaluate factors that affect FBW.

**Results:** Among the 94 fellow eyes of unilateral ERM; 71 eyes (75.5%) had a wide-based foveal pit. Both their FBW (418.9 ± 68.7 µm) and area of FAZ (0.38 ± 0.12 mm^2^) were significantly larger than those in the control group (323.2 ± 74.3 µm and 0.31± 0.17 mm^2^; *p* < 0.001 and *p* = 0.02, respectively), and they also had a thinner CFT and CST, and a wider RAT than the control group (*p* < 0.05 for all). In the control group, females had a wider FBW, a thinner CFT and CST, and a wider RAT than males (p < 0.05 for all). Regression analysis showed that female, a thinner CST and a larger FAZ were all associated with a larger FBW.

**Conclusion:** Fellow eyes of the unilateral ERM had a larger FBW and FAZ, a thinner CFT and CST, and a wider RAT than the normal population. This implicates that centrifugal macular traction may result in the formation of idiopathic ERM. Females had a wider FBW, a thinner CST, and a wider RAT than males, which may explain the higher prevalence of idiopathic ERM in females.

## Introduction

Epiretinal membrane (ERM) has a range of clinical presentations, from the milder cellophane maculopathy to the more severe macula pucker. With similar clinical findings and treatment options, the underlying etiologies could be secondary to retinal pathologies or idiopathic. ^1^ Idiopathic ERM forms with the migration of cells and collagen deposition either along the posterior hyaloid surface or from the clefts of the internal limiting membrane (ILM), the innermost layer of the retina.^2-4^ ILM is composed of interweaved footplate of the Muller cells, and natural ILM pores were found near the large vessels and towards the periphery of the normal retina. Clefts and deficits has been noticed in sections of the central retina where ERM coexists.^5^ It has been proposed that these ILM clefts in the macula may be due to the separation of the posterior hyaloid membrane or by the tangential tractional forces exerted on the central macula.^6 7^

In our previous study,^8^ a specific pattern of the fovea, a wide-based foveal pit was noted. Eyes with a wide-based foveal pit either had a similar wide-based foveal pit or vitreomacular diseases including ERM and the macular hole in their fellow eyes. The observed symmetry of foveal pit structure in bilateral eyes correspond to some previous study which demonstrated high consistency between bilateral foveal pits despite interpersonal variations in foveal pit parameters.^9^ However, the high prevalence of vitreomacular diseases in the fellow eyes suggests that such foveal contour should be related to some vitreomacular interface pathology. Among the vitreomacular diseases in the fellow eyes, ERM is the most prevalent; therefore, we propose that widening of the foveal base could be a result of tangential traction at the central macula and serve as one risk factor for ERM formation. To confirm this hypothesis, the foveal pit morphology as well as the macular structure of presumed normal fellow eyes of unilateral ERM were analyzed and compared with normal controls in this study.

## Methods

### Study Subjects

Patients with unilateral idiopathic ERM diagnosed between November 2016 and August 2019 in National Taiwan University Hospital were retrospectively collected, and their normal fellow eyes were enrolled as the study group. During the same period, patients who received preoperative OCT examination for cataract or refractive surgery were screened. Those who had regular, smooth foveal contour and no obvious maculopathy in both eyes were enrolled as the control group. All eyes with posterior staphyloma, previous retinal detachment, retinopathy of prematurity, retinal dystrophies, or retinal degenerations were excluded. Demographic data and ocular examinations including axial length, fundus photography, and OCT were collected. Axial length was measured with optical biometry (LENSTAR LS700, Haag-Streit, USA). Macular structure characteristics were measured on B-scan OCT and en face OCT angiography using Optovue Avanti RTVue XR OCT (Optovue, Inc., Fremont, CA, USA). The fundus images were obtained using either color fundus photography, fundus auto-fluorescence, or infrared image. Cases with incomplete data were excluded from this study. Finally, 94 fellow eyes of unilateral ERM were recruited in the study group, and 225 normal eyes were recruited in the control group. Among the 225 normal eyes, 94 of them who were gender and age-matched (same sex, age difference ≤ 5 years) to the cases in the study group were chosen for further paired comparison. This study was approved by the Ethics Committee and Institutional Review Board of National Taiwan University Hospital and adhered to the Declaration of Helsinki.

### Parameters of Macular Structure Characteristics

The parameters of macular structure characteristics included the foveal base width (FBW), central foveolar thickness (CFT), central subfield thickness (CST), central subfield volume (CSV), foveal avascular zone (FAZ), and retinal artery trajectory (RAT). The FBW was measured at a line that is 10 µm above the lowest point of the pit and parallel to the underlying RPE layer in the B scan OCT image. Both the horizontal and vertical sections were used to measure the width of the foveal base and the mean value was recorded as the FBW (Figure 1). The CFT was measured from the lowest point of the pit to the base of the retinal pigment epithelium layer. CST and CSV were measured at the central 1mm-diameter area around the foveal pit; CST was the mean retinal thickness in this area, and CSV was the volume of the retina under this area. FAZ was measured according to the en face OCT angiography. CST, CSV, and FAZ were all acquired automatically through the software of Optovue Avanti RTVue, and the centration was checked manually for precision. The technique for measuring RAT was modified according to the method proposed by Yoshihara et al.^6^ In short, the fundus images were rotated 90 degrees clockwise for right eyes and 90 degrees counterclockwise for left eyes using ImageJ (ImageJ version 1.47, National Institutes of Health, Bethesda, MD; available at http://imagej.nih.gov/ij/). The arcade arteries were manually dotted, with the first being the site where the retinal artery emanating from the optic disc. Twenty dots were labeled in each fundus photograph and the x-y coordination was generated with the “invert Y coordinate” function of ImageJ. Those coordinates were then fitted to the best fit curve with second degree polynomial equation (Figure 2).

**Figure 1.**
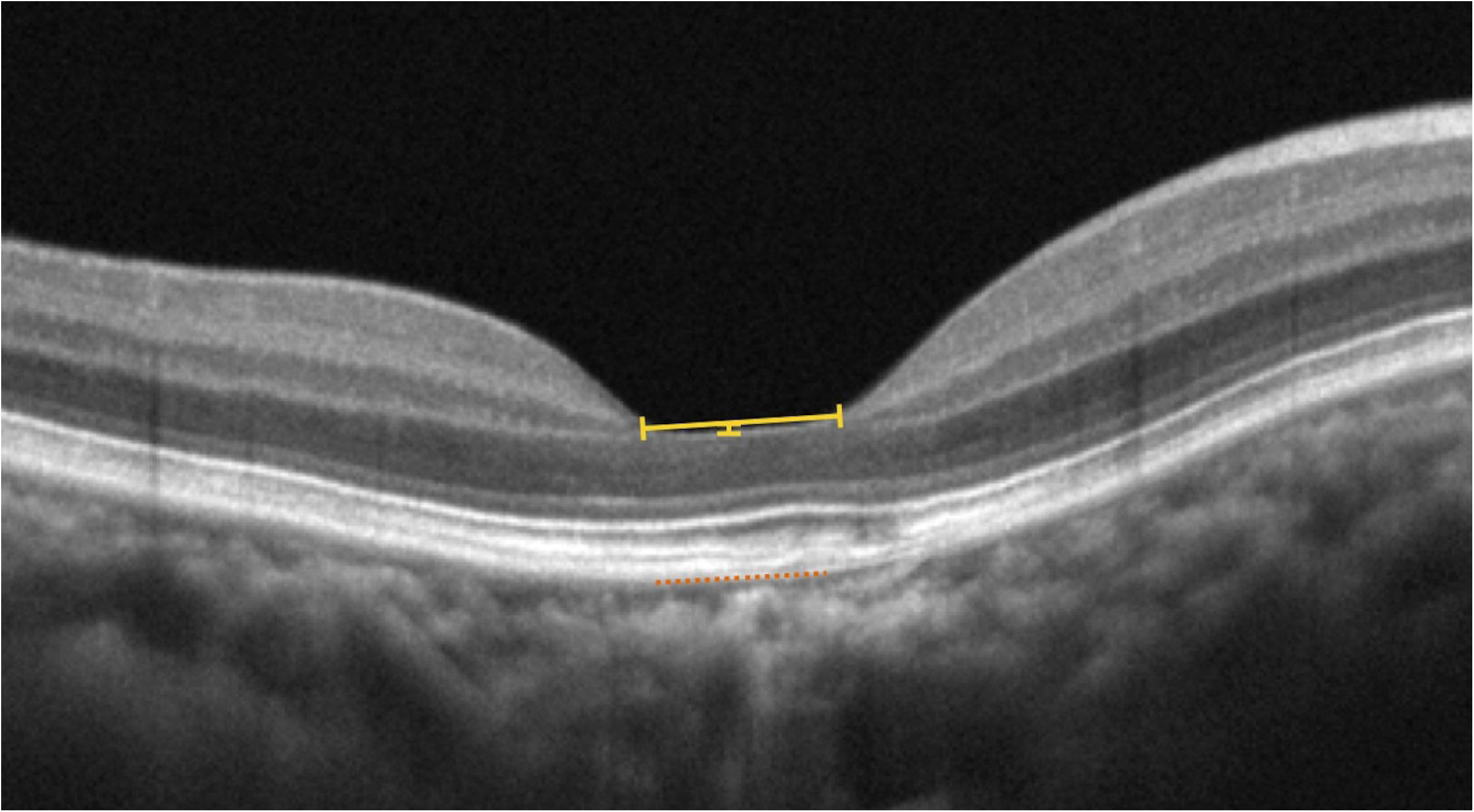
Foveal base width (horizontal yellow line) was measured at a line 10 µm above the lowest point of the foveal pit (vertical short yellow segment) and parallel to the retinal pigment epithelium layer (dotted line). This figure is a case of a wide-based foveal pit.

**Figure 2.**
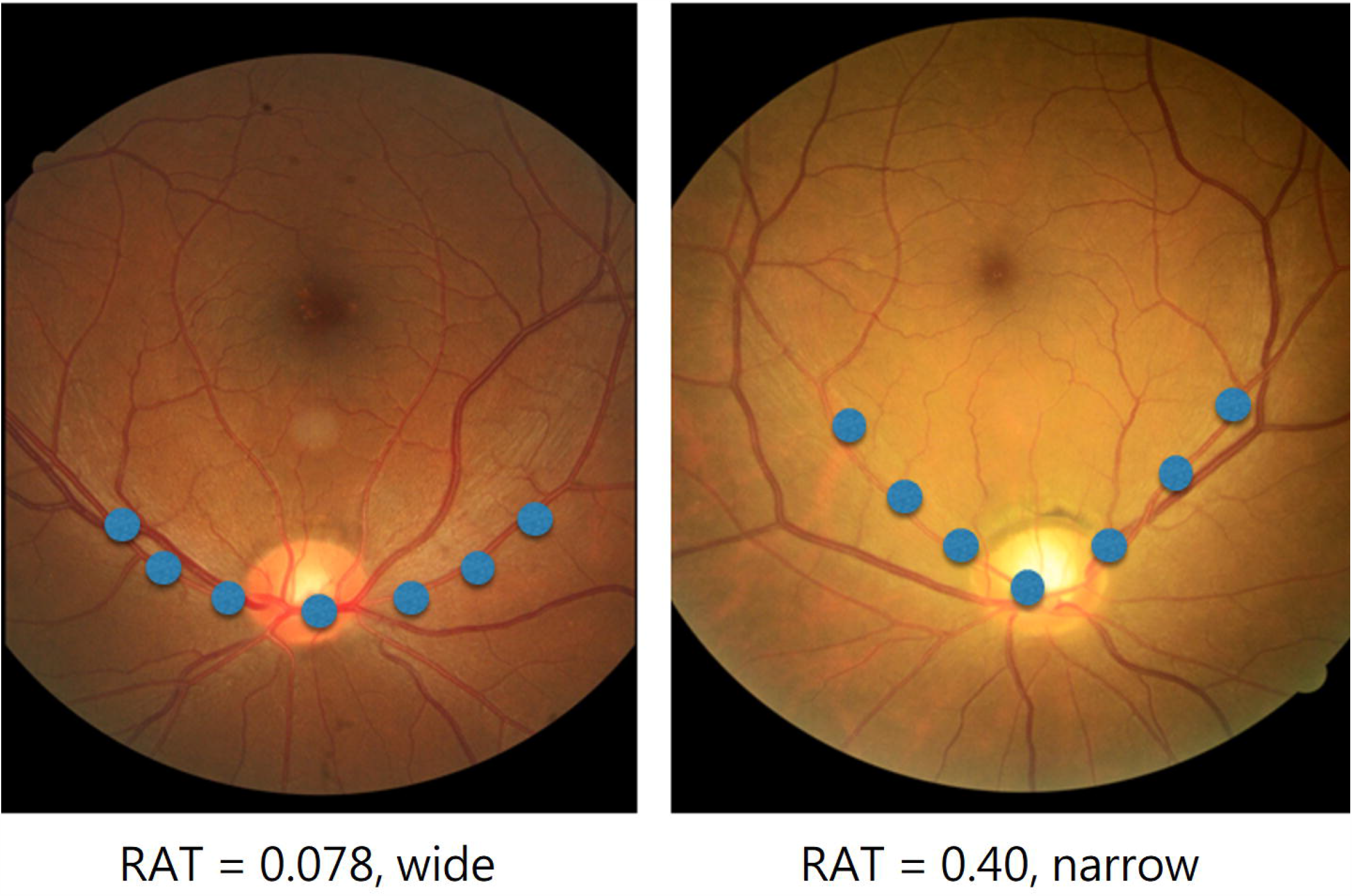
Measurement of the retinal artery trajectory (RAT) from color fundus photography. We manually placed 20 dots along the arcade arteries, with the first at the site where the retinal artery emanates from the optic disc (sample blue dots shown). These dots then fitted to the best fit curve with a second-degree polynomial equation using the “invert Y coordinate” function of ImageJ. The coefficient of the second-degree polynomial represents the width of the curve, with wide curves having smaller coefficients (left), and narrow curves having larger coefficients (right).

### Statistical Analysis

Student *t*-tests were used for comparison of parameters (axial length, FBW, CFT, CST, CSV, FAZ, and RAT) between different groups. A receiver operating characteristic (ROC) curve was plotted using FBW to predict if the fellow eye has ERM, and the area under the curve (AUC) was calculated. The data were analyzed using SPSS software (SPSS 22.0; SPSS Inc., Chicago, IL, USA). A *p*-value of less than 0.05 was considered statistically significant.

## Results

A total of 225 normal eyes and 94 fellow eyes of unilateral ERM were recruited for the study. The distributions of FBW in both groups are shown in Figure 3. For the normal population, most of the FBW was between 220 to 400 μm, while the distribution skews to the right with some FBW as large as near 600 μm. As for the fellow eyes of unilateral ERM, although the whole distribution ranges from 200 to 600 μm, most cases fall between 400 to 460 μm. Among the 225 normal eyes, the mean FBW was 323 μm, and the standard deviation (SD) was 74 μm. Hence, the mean plus one SD, which was 397 μm, was chosen as the cut-off point for the definition of wide-based foveal pit in the study.

**Figure 3.**
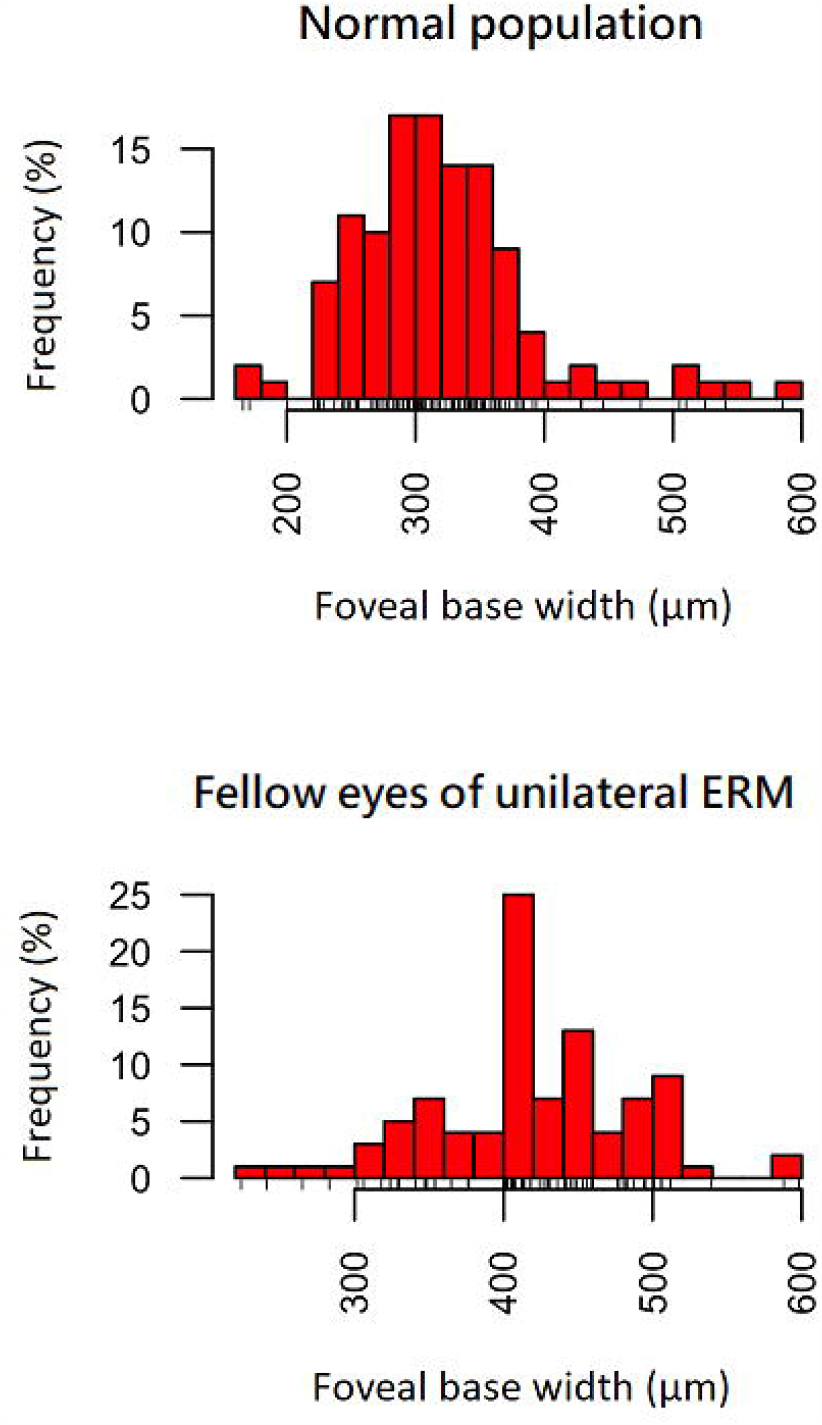
Histograms showing the distributions of foveal base width distribution in the normal population and in the fellow eyes of unilateral epiretinal membrane.

Among the 94 eyes in the study group, there were 58 female and 36 male patients. The average age was 66.9 ±7.2 years, and the mean axial length was 23.98 ± 1.24 mm. Compared to the age and gender-matched control group, the study group had a wider FBW (418.9 ± 68.6 µm vs. 323.2 ± 74.3 µm, p < 0.001), a thinner CFT (203.9 ± 20.0 µm vs. 209.6 ± 22.9 µm, p = 0.04), and a thinner CST (237.9 ± 19.7 µm vs. 248.7 ± 15.4 µm, p =0.001). 75.5% of eyes in the study group were recognized as having a wide-based foveal pit, in contrast to 9.8% in the control group (p < 0.001) (Table 1).

**Table. 1.** Comparison of Demographic data and Foveal Pit Characteristics between Fellow eyes of Unilateral Epiretinal Membrane patients and Age-Matched Normal Subjects.

|  | Fellow eyes of Unilateral Epiretinal Membrane (n = 94) |  |  | Age and Sex-Matched Normal Controls (n = 94) |  |  | p-value |
| --- | --- | --- | --- | --- | --- | --- | --- |
| AxL | 23.98 | ± | 1.24 | 23.49 | ± | 0.84 | 0.06 |
| FBW | 418.9 | ± | 68.6 | 323.2 | ± | 74.3 | <0.001 |
| CFT | 203.9 | ± | 20.0 | 209.6 | ± | 22.9 | 0.04 |
| CSV | 0.202 | ± | 0.017 | 0.198 | ± | 0.020 | 0.11 |
| CST | 237.9 | ± | 19.7 | 248.7 | ± | 15.4 | 0.001 |
| FAZ | 0.38 | ± | 0.12 | 0.31 | ± | 0.17 | 0.02 |
| RAT | 0.25 | ± | 0.15 | 0.35 | ± | 0.12 | <0.001 |

AxL: axial length (mm), CFT: central foveal thickness (μm), CST: central subfield thickness (μm), CSV: central subfield volume (μm<sup>3</sup>), FAZ: foveal avascular zone, FBW: foveal base width (μm), RAT: retinal artery trajectory.

### Gender Difference in Macular Structure Characteristics

Table 2 shows the differences in macular structure characteristics between male and female in the normal population as well as in the study group. In normal population, female had a wider FBW (335.7 ± 79.8 µm vs. 307.6 ± 62.0 µm, p =0.004), a thinner CFT (203.1 ± 19.3 µm vs. 221.8 ± 20.7 µm, p <0.001) and a thinner CST (245.3 ± 14.86 µm vs. 261.0 ± 18.3 µm, p = 0.049) than male. As for the study group, female also had a wider FBW (430.9 ± 60.6 µm vs. 397.2 ± 75.7 µm, p =0.03) and a thinner CST (234.3 ± 19.1 µm vs. 245.0 ± 19.4 µm, p =0.03) than male, but the difference in CFT was not statistically significant (202.2 ± 18.7 µm vs. 206.2 ± 21.6 µm, p = 0.37). Female had a wider RAT than male in the normal population (0.33 ± 0.11 vs. 0.40 ± 0.12, p = 0.011), while the RAT of the male in the study group (0.22 ± 0.14) was not only wider than male in the normal population but also wider than female in the study group (0.27 ± 0.16), although not statistically significant (p = 0.13).

**Table. 2.**
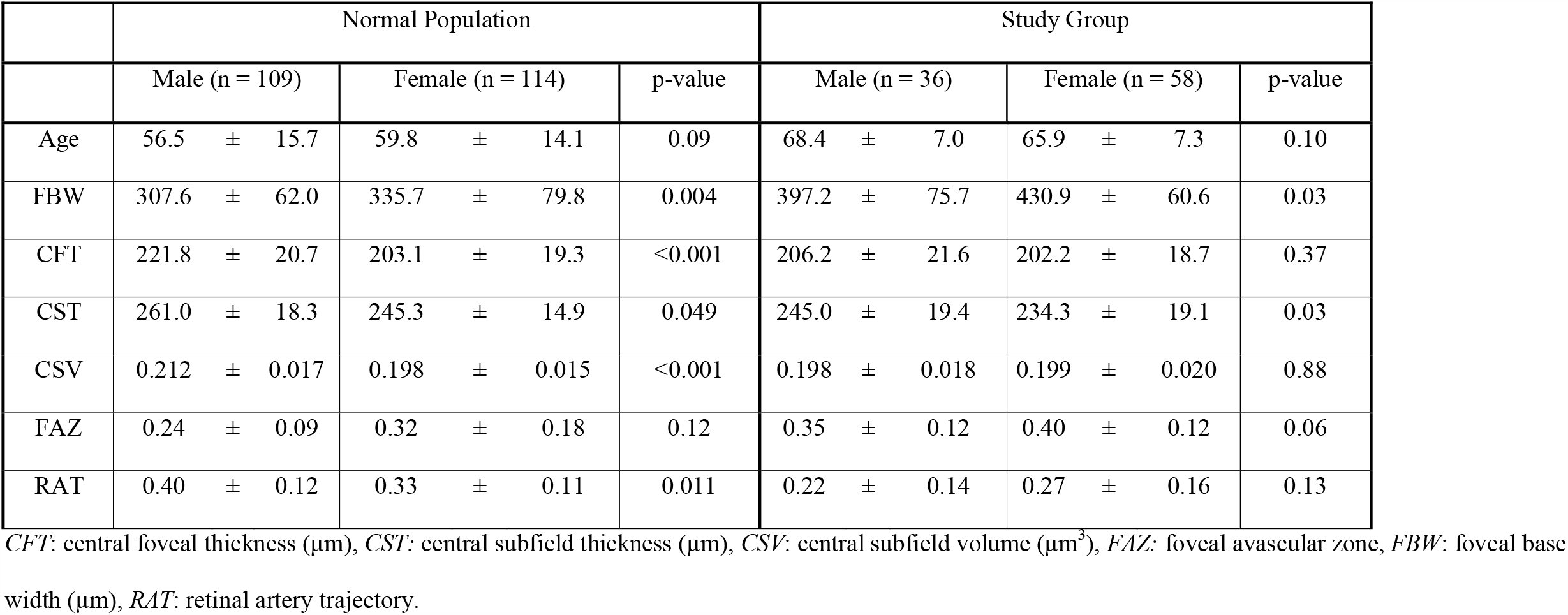
Comparison of macular structure parameters between male and female in normal controls and in the study group.

### Factors Associated with Foveal Base Width

Regression analysis was performed to evaluate the correlating factors for FBW. In the normal population, the FBW was inversely correlated with CFT, CST, and CSV, and positively correlated with FAZ (*p* < 0.05 for all). There was a strong correlation between female and a wider FBW (*p*=0.004). In the study group, similar results were observed with FBW negatively correlated with CST (*p* < 0.001) and positively correlated with FAZ (*p* < 0.001). Females also tended to have a wider FBW (*p* = 0.019). The FBW was not correlated with age, axial length, RAT, or PVD status in either group. (Table 3).

**Table 3.** Liner regression analysis for correlation with foveal base width in normal population and in the study group.

| Variables | Normal Population |  | Study Group |  |
| --- | --- | --- | --- | --- |
|  | Coefficient | p value | coefficient | p value |
| Age | 0.23 | 0.50 | -0.40 | 0.68 |
| Sex | -28.10 | 0.004 | -33.70 | 0.019 |
| AxL | -16.10 | 0.25 | -14.59 | 0.13 |
| PVD | 13.38 | 0.58 | 17.29 | 0.57 |
| CFT | -0.86 | <0.001 | -0.59 | 0.098 |
| CST | -1.879 | <0.001 | -1.806 | <0.001 |
| CSV | -2325.82 | <0.001 | -713.65 | 0.050 |
| FAZ | 283.73 | <0.001 | 291.21 | <0.001 |
| RAT | 37.94 | 0.52 | -6.68 | 0.89 |

*AxL*: Axial length, *CFT*: central foveal thickness ( $\mu\text{m}$ ), *CST*: central subfield thickness ( $\mu\text{m}$ ), *CSV*: central subfield volume ( $\mu\text{m}^3$ ), *FAZ*: foveal avascular zone,
*PVD*: posterior vitreous detachment, *RAT*: retinal artery trajectory.

### Predicting Epiretinal Membrane with Foveal Base Width of the Fellow Eyes

An ROC curve was plotted using FBW to predict if the fellow eye has ERM, and the AUC was 0.849. The best threshold was at 397 µm, with a sensitivity of 75.8% and specificity of 90.2%. (Figure 4)

**Figure 4.**
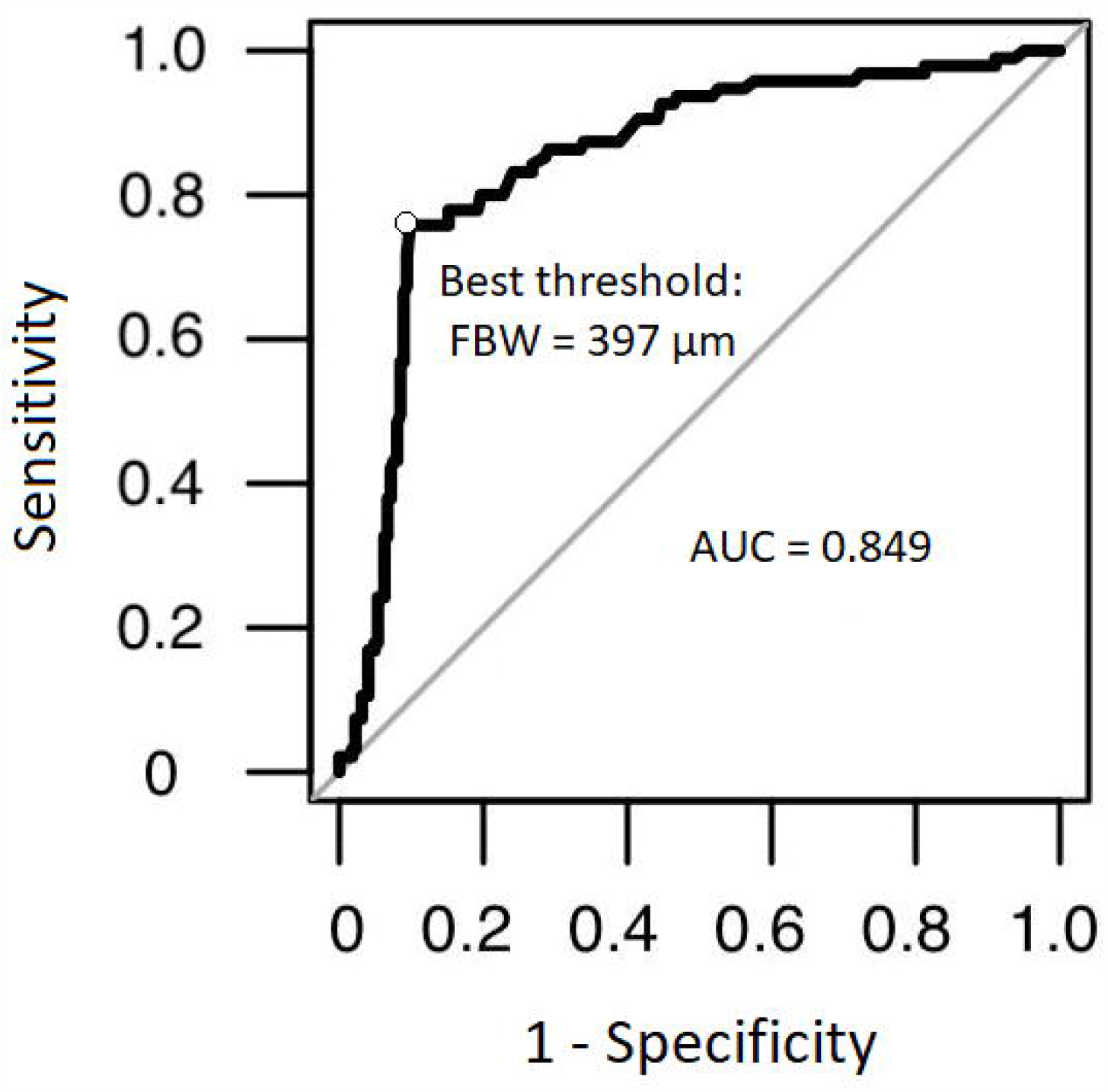
Receiver operation characteristic curve for predicting the presence of idiopathic epiretinal membrane using the foveal base width of the fellow eye.

## Discussion

Fovea development is visible at 22-weeks of gestational age and continues its maturing process into infants.^10^ The foveal depression continues to deepen after birth until 15 months, and it is formed by a vertical contraction of the central Muller cells.^11 12^ Afterwards, Muller cell processes contract horizontally, photoreceptors became more compact centrally, and these give rise to the base and the slope of the foveal pit. Under normal development, the fovea matures symmetrically in bilateral eyes; this gives rise to the design of using the fellow eyes of unilateral ERM patients as the study group to find the possible macular structure-related factors for the formation of idiopathic ERM in this study.

In this study, it was found that the fellow eyes of unilateral ERM have a wider foveal base and a larger FAZ than the normal population. Not only was the average FBW much wider, but more than three-fourth of them had a wide-based foveal pit, which is wider than the mean value plus one standard deviation of the normal population. This finding corresponds to the proposal that the retracting force of astrocytes in central fovea widens the foveal base, and such centrifugal force may result in internal limiting membrane clefts at the central macula, which may stimulate glial cell proliferation and collagen deposition, and eventually lead to ERM formation.^3,6^

From the standpoint of macular structure, while astrocytes contract and exert centrifugal force on the central macula, not only the force would widen the foveal pit but also would make it thinner. Thus, other anatomical parameters of the foveal pit might be influenced as well. This is compatible with our findings that CFT, CST, and CSV were all smaller in the study group than in normal controls. The results from the regression analysis also confirmed our assumption. The positive correlation of FAZ and FBW suggests that the widening of the foveal pit comes from outstretching of the fovea, not from pure thinning or atrophy of parafoveolar tissues. Furthermore, CFT, CST, and CSV were all negatively associated with FBW. Although CFT, CST, and CSV are three parameters representing similar concepts; there are still some differences among them. CFT is the thickness at the center of the foveola, while CST represents the average thickness of the central 1mm-diameter area. The negative correlation between FBW and CST/CFT suggests that the widening of the foveal base is due to the thinning of the parafoveaolar area, rather than the thickening of the central foveola. Such correlation is compatible with our hypothesis that some centrifugal force results in both widening of the foveal pit and thinning of central foveola.

If a centrifugal force does exist at the macula, theoretically the RAT will be widened. The results did show that the RAT was much wider in the fellow eyes of unilateral ERM patients than normal controls (p < 0.001). However, the regression analysis did not show a significant correlation between FBW and RAT. It is possible that the RAT varies a lot among the normal population, so the correlation between FBW and RAT would be masked. As for other factors such as age and axial length, neither of them was correlated with FBW. Despite the fact that axial length poses an optical effect on image acquisition which may lead to minimize measurement error,^13^ the association of FBW is not correlated with the axial length within 21-26mm. The status of PVD also had no correlation with FBW, which is reasonable as vitreomacular traction in non-PVD eyes is anteroposterior traction, rather than tangential traction that could affect the FBW.

Gender difference in macular structure is a topic of interest. Previous studies have identified that the CST is thicker in male than in female.^9 14 15^ Our previous study also observed a female preponderance in eyes with a wide-based foveal pit.^8^ In this study, it was found that female not only had a wider FBW and a thinner CFT/CST, but also had a wider RAT. This is compatible with the assumption that females may have a stronger centrifugal force at the macula that results in this phenomenon. Such result could also explain the finding by Poplin et al.^16^ that the “hot spot” for gender differentiation by deep learning models focused on retinal arteries in fundus photography. Furthermore, these observed gender differences in macular structure could also explain the female preponderance in ERM and macular hole formation.^17-19^ Interestingly, males who had an idiopathic ERM had the widest RAT in their fellow eyes among all in this study; this means that a really strong centrifugal traction force is needed for male to develop idiopathic ERM.

In conclusion, this study found that the fellow eyes of unilateral ERM had a larger FBW and FAZ, a thinner CFT and CST, and a wider RAT than the normal population. This implicates that centrifugal macular traction may result in the formation of idiopathic ERM. We proposes that the centrifugal traction force at macula leads to widening of the foveal pit and thinning of central foveola, and it eventually may produce some cracks in ILM which harbor retinal glial cell proliferation and ERM formation. Female was found to have a wider FBW, a thinner CST and CST, and a wider RAT than male, which may explain the higher prevalence of idiopathic ERM in the female.

## Data Availability

Data are available from the authors upon reasonable request.

## Abbreviations

CFT: central foveolar thickness
CST: central subfield thickness
ERM: epiretinal membrane
FAZ: foveal avascular zone
OCT: optical coherence tomography
RAT: retinal artery trajectory.

## References

1. Snead DR, James S, Snead MP. Pathological changes in the vitreoretinal junction 1: epiretinal membrane formation. Eye (Lond). 2008;22(10):1310–7. doi: 10.1038/eye.2008.36

2. Greven CM, Slusher MM, Weaver RG. Epiretinal membrane release and posterior vitreous detachment. Ophthalmology. 1988;95(7):902–5.

3. Bovey EH, Uffer S. Tearing and folding of the retinal internal limiting membrane associated with macular epiretinal membrane. Retina. 2008;28(3):433–40. doi: 10.1097/IAE.0b013e318150d6cf

4. Kishi S, Shimizu K. Oval defect in detached posterior hyaloid membrane in idiopathic preretinal macular fibrosis. Am J Ophthalmol. 1994;118(4):451–6.

5. Wolter JR. Pores in the Internal Limiting Membrane of the Human Retina. Acta Ophthalmol (Copenh). 1964;42:971–4.

6. Yoshihara N, Sakamoto T, Yamashita T, et al. Wider retinal artery trajectories in eyes with macular hole than in fellow eyes of patients with unilateral idiopathic macular hole. PLoS One. 2015;10(4):e0122876. doi: 10.1371/journal.pone.0122876

7. Yamashita T, Asaoka R, Tanaka M, et al. Relationship between position of peak retinal nerve fiber layer thickness and retinal arteries on sectoral retinal nerve fiber layer thickness. Invest Ophthalmol Vis Sci. 2013;54(8):5481–8. doi: 10.1167/iovs.12-11008

8. Ma I-H, Yang C-M, Hsieh Y-T. Wide-based Foveal Pit: A Predisposition to Idiopathic Epiretinal Membrane. medRxiv. 2020:2020.08.13.20174334. doi: 10.1101/2020.08.13.20174334

9. Wagner-Schuman M, Dubis AM, Nordgren RN, et al. Race- and sex-related differences in retinal thickness and foveal pit morphology. Invest Ophthalmol Vis Sci. 2011;52(1):625–34. doi: 10.1167/iovs.10-5886

10. Hendrickson AE, Yuodelis C. The morphological development of the human fovea. Ophthalmology. 1984;91(6):603–12. doi: 10.1016/s0161-6420(84)34247-6

11. Yuodelis C, Hendrickson A. A qualitative and quantitative analysis of the human fovea during development. Vision Res. 1986;26(6):847–55.

12. Bringmann A, Syrbe S, Gorner K, et al. The primate fovea: Structure, function and development. Prog Retin Eye Res. 2018;66:49–84. doi: 10.1016/j.preteyeres.2018.03.006

13. Sampson DM, Gong P, An D, et al. Axial Length Variation Impacts on Superficial Retinal Vessel Density and Foveal Avascular Zone Area Measurements Using Optical Coherence Tomography Angiography. Invest Ophthalmol Vis Sci. 2017;58(7):3065–72. doi: 10.1167/iovs.17-21551

14. Wong AC, Chan CW, Hui SP. Relationship of gender, body mass index, and axial length with central retinal thickness using optical coherence tomography. Eye (Lond). 2005;19(3):292–7. doi: 10.1038/sj.eye.6701466 [published Online First: 2004/07/20]

15. Kashani AH, Zimmer-Galler IE, Shah SM, et al. Retinal thickness analysis by race, gender, and age using Stratus OCT. Am J Ophthalmol. 2010;149(3):496–502 e1. doi: 10.1016/j.ajo.2009.09.025 [published Online First: 2010/01/01]

16. Poplin R, Varadarajan AV, Blumer K, et al. Prediction of cardiovascular risk factors from retinal fundus photographs via deep learning. Nat Biomed Eng. 2018;2(3):158–64. doi: 10.1038/s41551-018-0195-0 [published Online First: 2019/04/25]

17. Duan XR, Liang YB, Friedman DS, et al. Prevalence and associations of epiretinal membranes in a rural Chinese adult population: the Handan Eye Study. Invest Ophthalmol Vis Sci. 2009;50(5):2018–23. doi: 10.1167/iovs.08-2624

18. Fraser-Bell S, Ying-Lai M, Klein R, et al. Prevalence and associations of epiretinal membranes in latinos: the Los Angeles Latino Eye Study. Invest Ophthalmol Vis Sci. 2004;45(6):1732–6.

19. Miyazaki M, Nakamura H, Kubo M, et al. Prevalence and risk factors for epiretinal membranes in a Japanese population: the Hisayama study. Graefes Arch Clin Exp Ophthalmol. 2003;241(8):642–6. doi: 10.1007/s00417-003-0723-8

